# Air filtration mitigates aerosol levels both during and after OGD procedures

**DOI:** 10.1101/2022.08.23.22279118

**Authors:** Frank Phillips, Jane Crowley, Samantha Warburton, Adolfo Parra-Blanco, George S.D. Gordon

**Affiliations:** NIHR Nottingham Biomedical Research Centre, Nottingham University Hospitals NHS Trust and the University of Nottingham, Nottingham, UK; Faculty of Engineering, University of Nottingham, Nottingham, UK

**Author notes:** Contribution: study concept and design, acquisition of data, drafting of manuscript, critical revision of manuscript. Contribution: collected and curated data. Contribution: study concept and design, obtained funding, acquisition of data, critical revision of manuscript, study supervision. Contribution: study concept and design, statistical analysis, drafting of manuscript, critical revision of manuscript, study supervision. **FUNDING** The authors also thank Nottingham University Hospitals NHS Trust for funding the rental of a particle counter, Norgine Pharmaceuticals for sponsoring the purchase of a particle counter. GSDG would like to acknowledge a UKRI Future Leaders Fellowship (MR/T041951/1). **DATA AVAILABILITY STATEMENT** Data associated with this publication is available at http://dx.doi.org/10.17639/nott.7112 Code used for data analysis in this publication can be found at https://github.com/gsdgordon/aerosols.

**Keywords:** COVID-19, Endoscopy, Aerosol generating procedures, Air filtration

## Abstract

**Objectives:** Upper GI endoscopies are aerosol generating procedures (AGPs), increasing risk of spreading airborne pathogens. We aim to quantify mitigation of airborne particles via improved ventilation, specifically laminar flow theatres and portable HEPA filters, during and after upper GI endoscopies.

**Methods:** This observational study included patients undergoing routine oral gastroscopy in a standard endoscopy room with 15-17 air changes per hour, a standard endoscopy room with portable HEPA filtration unit, and a laminar flow theatre with 300 air changes per hour. A particle counter (diameter range 0.3µm-25µm) took measurements 10cm from the mouth. Three analyses were performed: whole procedure particle counts, event-based counts and air clearance estimation using post-procedure counts.

**Results:** Compared to a standard endoscopy room, for whole procedures we observe a 28.5x reduction in particle counts in laminar flow (p<0.001) but no significant effect of HEPA filtration (p=0.50). For event analysis we observe for lateral flow theatres reduction in particles >5µm for oral extubation (12.2x, p<0.01), reduction in particles <5µm for coughing/gagging (6.9x, p<0.05) and reduction for all sizes in anaesthetic throat spray (8.4x, p<0.01) but no significant effect of HEPA filtration. However, we find that in the fallow period between procedures HEPA filtration reduces particle clearance times by 40%.

**Conclusions:** Laminar flow theatres are highly effective at dispersing aerosols immediately after production and should be considered for high-risk cases where patients are actively infectious or supply of PPE is limited. Portable HEPA filers can safely reduce fallow time between procedures by 40%.

## Introduction

Prior to the COVID-19 pandemic, gastrointestinal endoscopy was not considered an Aerosol Generating Procedure (AGP). However, it has now been proven that upper and lower endoscopic procedures can generate aerosols, although their infectivity by SARS-Cov-2 remains unclear, especially for lower GI endoscopy (1,2). There have been various approaches to mitigating aerosols during endoscopy, including the use of facemasks on patients which is increasingly being adopted clinically (3,4) and the substitution of per-oral for trans-nasal endoscopy which can reduce aerosol production by 50% (1). Improved is another key aerosol mitigation strategy but recommendations vary widely: newly designed endoscopy rooms in the UK require at least 10 air changes per hour (ACH), and negative pressure (5). Operating theatres with laminar air flow are widely used as means of limiting airborne transmission of pathogens and contaminants (6): for example, they have been shown to reduce aerosol concentration by factors of 100 or more in arthroplasty (7). Laminar flow theatres are therefore a promising approach to mitigate aerosols during digestive endoscopy. Gregson et al. measured particle counts during upper digestive endoscopy in an operating theatre with laminar flow but during measurements the ventilation was set to ‘standby’, reducing the ACH from 500-600 to 25 (8). Previous work has shown that reducing ACH to 25 does not significantly reduce particles measured from volitional coughing, but reducing ACH to zero dramatically increases particles (9). However, no comparison has been performed with ventilation conditions in typical endoscopy rooms without laminar flow and with ACH typically lower than a laminar flow theatre on standby, nor have typical events encountered in endoscopy (involuntary gagging, extubation, throat spray). The mitigating effect of fully operative laminar flow on aerosol levels during digestive endoscopy compared to typical endoscopy rooms has therefore not been assessed.

Portable high-efficiency particulate air (HEPA) filtration units are a lower-cost alternative to laminar flow systems for mitigating aerosols. Numerous studies have shown the ability of a range of portable HEPA units to remove aerosols from the air (10), but these have mostly been conducted under controlled laboratory conditions. More recently, clinical studies in intensive care units showed that portable HEPA filtration units can significantly reduce viable SARS-CoV-2 in air samples (11,12). The use of portable HEPA filtration has been proposed for use in endoscopy units (13) but there have been no clinical studies on the effect under typical procedure conditions.

## Methods

The methodology used for this study is based on that we developed for a previous ‘baseline’ study of aerosol generation in digestive endoscopy (1). This is a prospective observational study. Health Research Authority and ethical approval was granted by the Wales Research Ethics Committee prior to the start of the study (IRAS no. 285595). We included patients undergoing routine upper GI endoscopy on the lists of thirteen different participating endoscopists at the Endoscopy unit of the Nottingham University Hospitals NHS Trust Treatment Centre between October 2020-March 2021.

The inclusion criteria were adult patients >18 years with capacity to consent. Procedures were performed as they normally would be in clinical practice. Patients chose whether they wanted sedation and procedures were performed with CO2 or air for insufflation and intermittent suctioning.

### Experimental procedure

We measured the concentration of aerosols (<5µm diameter) and droplets (>5µm diameter) produced during typical upper gastrointestinal endoscopy procedures conducted both in standard endoscopy rooms (n=33), standard endoscopy rooms with portable HEPA filtration units (Air Sentry Limited, Wiltshire, UK; n=4) and in laminar flow theatres (n=4 full procedures, n=9 for volitional cough and throat spray). These 3 different ventilation scenarios and typical airflow patterns are shown in Figure 1a,b, and c respectively. Ventilation effectiveness is affected by room size, placement of air filtration units and placement of particle counters. For reasons of practicality, we here consider a limited subset of these possible variations that represents a typical clinical scenario. Specifically, all room used are in the same endoscopy unit with similar ventilation conditions (in terms of ACH). For the arm of the study with the portable HEPA filtration unit, only a single room was available but the unit was professionally installed by the supplier and so is assumed to be operating to a standard expected for a wider deployment. The laminar flow theatres are, by necessity, somewhat different to conventional endoscopy rooms. However, this is representative of a practical mitigation strategy, which is what we primarily seek to examine.

**Figure 1.**
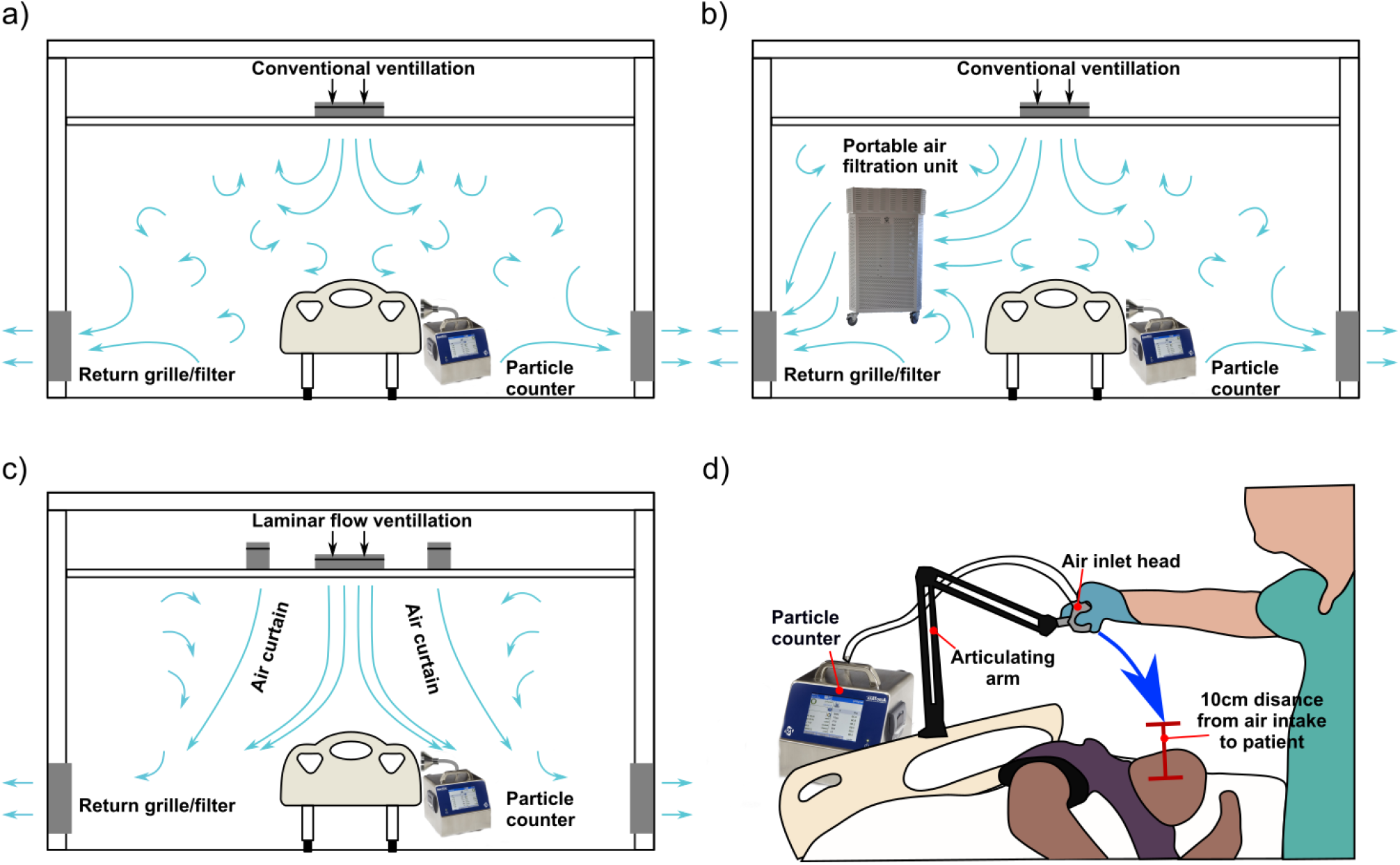
Schematic showing different ventilation conditions and measurement set-up: a) conventional ventilation with typically weaker flow and re-circulating air currents, b) conventional ventilation with portable HEPA filtration unit acting to increase the rate of air filtration and slightly increase air flow, c) laminar flow ventilation which creates powerful curtains of air to quickly remove all particles creating an ultra-clean environment, d) diagram showing typical measurement configuration with air-inlet to particle counter place on articulating arm and lowered to within 10cm of the patient’s mouth.

In the standard endoscopy rooms, the nominal ACH rate is 15-17 while for the laminar flow theatre it is 300. This is averaged across several rooms, but is measured by the in-house team using a balometer: the air inflow rate is measured in m^3^/hour and is then divided by the room volume in m^3^. The endoscopy rooms used were within the same endoscopy suite with similar ventilation, size, air temperature and humidity levels. All present in the room wore enhanced PPE which minimised the contribution of additional human aerosol sources. All procedures considered are OGD procedures, which we previously showed were AGP (1).

Particle counts were measured and analysed using an AeroTrak portable particle counter (TSI, Shoreview MN, model 9500-01) with inlet tube place 10cm from the patient’s mouth (methodology described in (1)), shown in Figure 1d. Previous studies have shown that detecting particles at distances greater than 10cm from source dramatically reduces detectable particle count (requiring ultra-clean rooms) and biases the size of particles detected (9). Therefore, to maximise detection of relevant particle concentrations produced by patients above relatively high background particle level and for comparability with previous studies (2) a distance of 10cm is chosen. The patient’s mouth is considered the main source of potentially infectious particles, since all other persons in the room are wearing FFP3-grade (N95) masks for this study.

We compared aerosol and droplet concentrations produced from whole procedures (median duration of 7.2 minutes), but we normalise counts to a 20 minute procedure by multiplying total particle count by the appropriate ratio. This to enable comparison between different types of procedures, such as colonoscopy. Sedation conditions vary across patients from only using Xylocaine throat spray to use of Midazolam. However, previous studies including our own have not found significant effect on aerosol production (1,14). We also analyse data from aerosol-producing events using a background subtraction approach described in our previous methodology (1). Specifically, we consider the following individual aerosol generating events: oral extubation, coughing/gagging during procedure, anaesthetic throat spray and volitional cough.

For the analysis of the inter-procedure particle counts, there was a mixture of lower- and upper-GI procedures preceding the fallow period (44% lower GI, 56% upper GI). This may affect the concentration of particles at the start of the fallow time: upper GI procedures produce more particles per unit time, though this may be compensated by the shorter duration (< 3x) compared to lower GI procedures (1). To avoid this problem, for the inter-procedure periods we consider the rate of change of particle counts, i.e. the gradients with respect to time. Because the HEPA filtration unit is significantly less powerful than a laminar flow system, a gradient approach will also be able to more sensitively quantify its effect, analogous to our ‘causal difference’ approach to measuring aerosol generating events (1). First, we identified all periods when particle count was continuously decreasing (e.g. when no people are present in the room), which we denote as particle ‘sink’ windows, defined when particle concentration decreases for 4 or more consecutive measurements. Across 4 fallow periods measured in standard endoscopy rooms with portable HEPA filtrations units, we identified 490 such sink windows. As a control, we identified 2305 sink windows across 33 procedures in standard endoscopy rooms without a HEPA filter. Within these sink windows, we compute the rate of clearance of particles (i.e. the gradient) and fit an exponential model to determine a dispersal rate constant. We then extrapolate to estimate the time taken to clear 50% of particles in the room.

### Statistical analysis

All statistical analysis was performed using the MATLAB software package (The MathWorks Inc., Massachusetts). Building on existing models of aerosol production in the respiratory tract we use a log-normal distribution to model the distribution of total particle counts (15). For the whole procedure data, a logarithm of the data is first computed (to convert the data from log-normal to normal) and then a *t-*test is applied to compute *p*-values. The mean values and confidence intervals resulting from this analysis become ratios when the inverse logarithm (i.e. exponential) is applied to convert back to raw particle counts. For individual events the data distribution is modelled as the sum of a log-normal and normal distribution to account for negative values of particle counts that can arise from the subtraction step. A Monte-Carlo sampling method is therefore used to provide numerical estimates of *p*-value and numerically estimate mean ratios and confidence intervals between events (16).

*A priori* power calculations based on limited previous studies of particle size distribution in coughing and sneezing we determined that with around 5 replicates per patient we can detect an effect size (Cohen’s d) of 1.98, which is sufficient to differentiate between a cough and sneeze (17,18). We also conducted a retrospective power analysis to determine whether sample sizes are sufficient. This uses the measured mean and standard deviation of particle counts under tested ventilation conditions and compares this to the null hypothesis that they follow the same distribution as the standard ventilation conditions.

For the inter-procedure analysis, the distribution of slopes during ‘sink periods’ is expected to be non-normal (‘long tail’ behaviour is observed empirically) and so we apply a Mann-Whitney U test to establish statistical significance of any observed difference. By randomly sub-sampling the data (bootstrapping) and repeating we can also obtain confidence intervals and a power calculation for the slope analysis.

## Results

The relevant demographics of the patients used in the 3 different rooms are highly similar enabling a fair comparison (shown in Table 1), though we note that our previous work found that among these features, only the presence of a hiatus hernia seemed to impact aerosol production.

**Table 1.** Summary table showing demographic data for patients enrolled in this study.

| **Ventilation scenario**<br>**Variable** | **Conventional ventilation** | **Conventional ventilation + portable HEPA filter** | **Laminar flow theatre** |
| --- | --- | --- | --- |
| ***n*** | 33 | 4 | 4 |
| **Age** | *Range: 24-93*<br>*Median: 63* | *Range: 24-71*<br>*Median: 58* | *Range: 41-74*<br>*Median: 52* |
| **Sex** | *Male: 20, Female: 13* | *Male: 1, Female: 3* | *Male: 1, Female: 3* |
| **BMI** | *Range: 16.3-38.2*<br>*Median: 24.8* | *Range: 19.8-26.3*<br>*Median: 24.8* | *Range: 24.9-39.3*<br>*Median: 32.4* |
| **Smoking** | *Smoker: 8*<br>*Non-smoker: 24* | *Smoker: 1*<br>*Non-smoker: 3* | *Smoker: 1*<br>*Non-smoker: 3* |
| **Hiatus hernia** | *Yes: 9, No: 24* | *Yes: 1, No: 3* | *Yes: 1, No: 3* |
| **Sedation** | *Midazolam: 14*<br>*Throat spray only: 19* | *Midazolam: 2*<br>*Throat spray only: 2* | *Midazolam: 3*<br>*Throat spray only: 1* |

For the whole procedure analysis, summarised in Figure 2a, in the aerosol size range we find no significant reduction in total particle count using the HEPA filtration unit (p=0.50) but a significant reduction when using a laminar flow room compared to standard endoscopy room (28.5x, 95%CI:13.9-58.3, p<0.001) and to standard endoscopy room with portable HEPA filtration unit (37.5x, 95%CI:5.7-245.5, p<0.05). A similar trend is observed for droplets (>5µm diameter) with a significant reduction in count for laminar flow theatre compared to the standard endoscopy room (30.7x, 95%CI: 16.9-55.9, p<0.001), and standard endoscopy room with HEPA filtration unit (50.0x, 95%CI:10.8-231.4, p<0.001).

**Figure 2.**
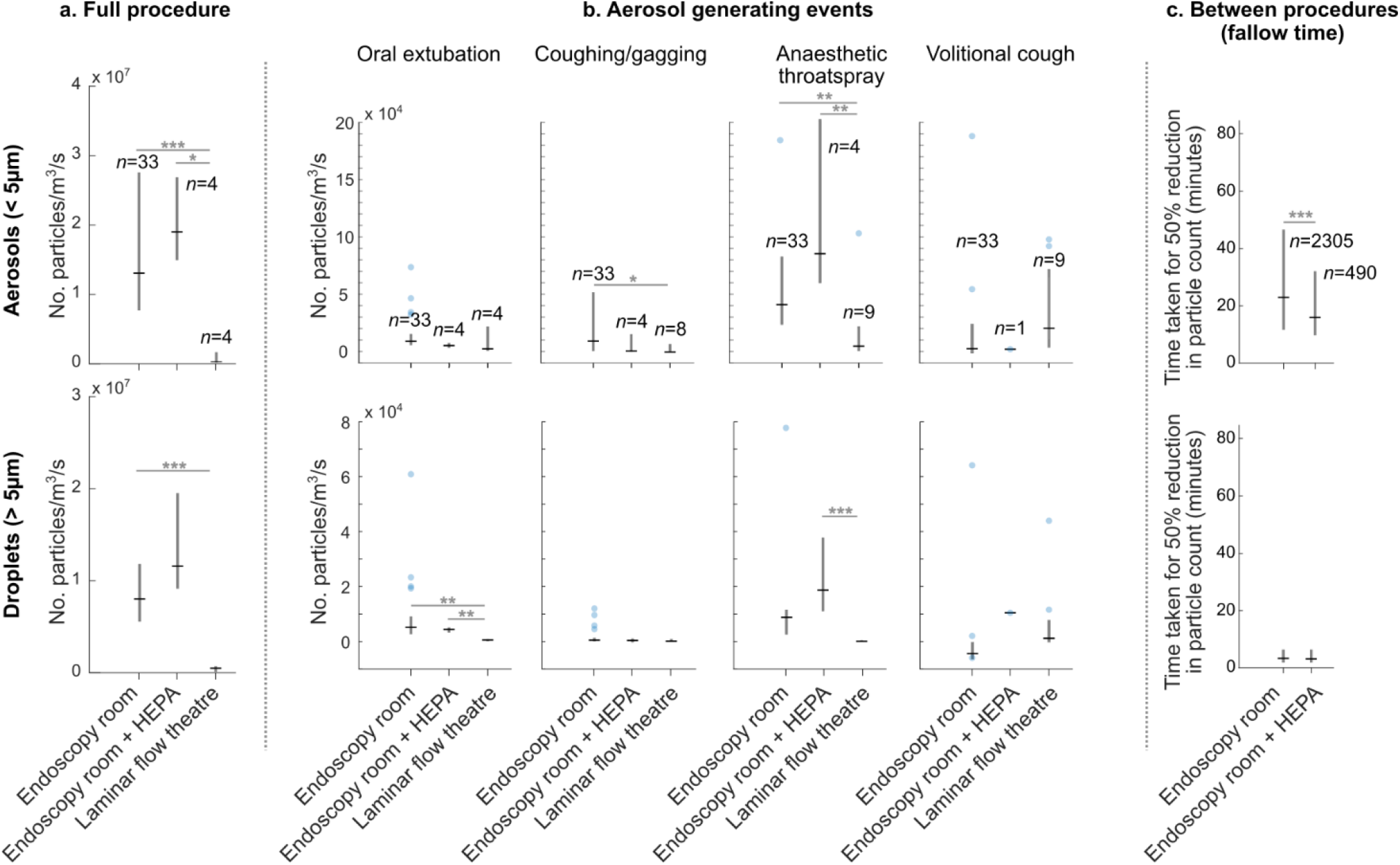
Effect of different room ventilation schemes on aerosol and droplet counts. a) Total particle counts across whole procedures. b) Comparison of 3 aerosol generating events. c) Investigation of particle clearance rate with and without portable HEPA filter, showing significant speed increase. *p<0.05, **p<0.01, ***p<0.001.

*A posteriori* analysis of study power suggests that, given the effect sizes estimated from measurements, to test the effect of the laminar flow theatre with power 0.9 and p=0.05 would need a sample size of n=3, and a with the measured sample size of n=4, study power is >0.999. To test the effect of the portable HEPA air filtration unit with power 0.9 and p=0.05 would require sample size n > 700, which would be challenging to measure given available numbers of patients. Our conclusion with our smaller sample of n=4 is therefore that the effect size of this mitigation strategy is very small, significantly less than for the laminar flow theatre.

We next consider individual aerosols generating events: oral extubation, coughing/gagging anaesthetic throat spray, and volitional cough (summarised in Figure 2b). For oral extubation, we find that in the aerosol size range particle counts are statistically comparable across the 3 room types, but in the droplet size range particle counts are significantly reduced in laminar flow room compared to standard endoscopy room (12.2x, 95%CI:5.0-38.3, p<0.01) and to standard endoscopy room with portable HEPA filtration (10.1x, 95%CI:4.0-35.7, p<0.01). The average particle diameter for oral extubation is significantly smaller in laminar flow (0.22*μ*m) compared to endoscopy room (2.8*μ*m, p<0.05) and to endoscopy room with a portable HEPA filter (4.6*μ*m, p<0.01). Together, these results suggest laminar flow removes larger particles very effectively, either through direct dispersal or evaporation, although we would not expect evaporation to have major impact over such short distances of travel (0.1m) particularly for larger particles (9). For coughing/gagging in the aerosol size range we measure a significant reduction in laminar flow theatres compared to endoscopy (6.9x, 95%CI:1.22-61.9,p<0.05) but find no significant difference compared to standard endoscopy room with portable HEPA filtration system. In the droplet size range, we find no significant difference between any of the room types, but this may simply reflect the small average particle size of coughing/gagging (1). For the application of anaesthetic throatspray in the aerosol size range we find significant reduction in laminar flow theatres compared to standard endoscopy rooms (8.4x, 95%CI: 2.03-64.1,p<0.01) and standard endoscopy rooms with portable HEPA filtration (20.7x, 95%CI: 2.9-199.3,p<0.01). A similar trend is observed in the droplet size range with laminar flow theatres measuring fewer particles than standard endoscopy rooms (46.0x, 95%CI:7.4-438.6,p<0.01) and standard endoscopy rooms with portable HEPA filtration (169.0x, 95%CI: 21.2-1855.3,p<0.001). For volitional coughing, we did not observe a significant reduction (p=0.11) between laminar flow and standard endoscopy rooms (comparison not possible with HEPA filtered room due to only one recorded event). We hypothesise this may be due to the substantially higher particle velocity for volitional coughing compared to less forceful involuntary gagging, enabling more airborne particles to reach the detector. This finding is consistent with previous work measuring volitional coughs in laminar flow theatres (9).

Finally, for our analysis of particle clearance rates (presented in Figure 2c.) we found that with the HEPA filtration units the median 50% clearance time was 16.8 minutes, compared to 23.8 minutes without HEPA filtration. Further, we found that the decay rate in the aerosol size range with the HEPA filter is 1.41x faster (p<0.001), implying an effective increase in air change rate from 15-17 to 21-24 ACH. We did not observe any significant reduction for particles in the droplet size range, likely because these particles clear much more quickly due to gravity (median: 3.1 minutes for 50% clearance). *A posteriori* analysis of this data shows that with our sample size of *n=490* and *p=0*.*05*, the study power is 0.997, giving a high degree of confidence to the results. For the laminar flow theatre, the particle counts between procedures are exactly zero due to very fast particle clearance, faster than can be detected using our present experiment.

## Discussion

Overall, we find that use of laminar flow theatres significantly reduces aerosols and droplets measured near the patient’s mouth, typically by a factor of >5x during upper GI endoscopic procedures. This finding also applies to individual aerosol generating events (oral extubation, coughing/gagging, application of throat spray), which are significantly reduced in magnitude (>5x).

However, respiratory coughing may still pose a risk as this is not significantly reduced. We do not find a significant reduction in particle counts during procedures using portable HEPA filtration units, implying their effect is too small to be measured with our sample size, particularly when used in a room with pre-existing adequate ventilation. This is expected because these filters are significantly less powerful than the whole-room ventilation in laminar flow theatres. However, by analysing fallow periods we find that portable HEPA filtration units can increases aerosol clearance speed by ∼40%, which could reduce safe fallow time between procedures by 5-7 minutes.

## Data Availability

Data associated with this publication is available at http://dx.doi.org/10.17639/nott.7112 Code used for data analysis in this publication can be found at https://github.com/gsdgordon/aerosols

## ACKNOWLEDGEMENTS

The authors thank Guru Aithal for critically reviewing the manuscript; Martin James and Bu Hayee for reviewing the study protocol; Matthew Sanderson, Andy Wragg, Nottingham University Hospitals Research and Innovation, University of Nottingham Biomedical Research Centre, Karren Staniforth, Laura Leman, Nina Duffy, Allison Ball and the Endoscopy Unit Staff in their support of the development of this study; the NIHR Aerosol Generating Procedures Group for their support during the study; Tina Rodriguez, Paul Brocklebank, Mirela Pana, Sabina Beg, Stefano Sansone, James Catton, Emilie Wilkes, Lorraine Clark, Andrew Horton, John White, Suresh Vasan Venkatachalapathy, Aida Jawhari, Ioannis Varmpompitis, and Muthuram Rajaram for performing endoscopic procedures in this study; and Olympus for loan of the trans-nasal endoscopes

## PATIENT CONSENT

Obtained

## ETHICS APPROVAL

Wales Research Ethics Committee

## REFERENCES

1. Phillips F, Crowley J, Warburton S, Gordon GSD, Parra-Blanco A. Aerosol and droplet generation in upper and lower GI endoscopy: whole procedure and event-based analysis. Gastrointest Endosc. 2022;96:603–11.

2. Chan SM, Ma TW, Chong MKC, Chan DL, Ng EKW, Chiu PWY. A proof of concept study: esophagogastroduodenoscopy is an aerosol-generating procedure and continuous oral suction during the procedure reduces the amount of aerosol generated. Gastroenterology. 2020;159:1949–51.

3. Bojórquez A, Larequi FJZ, Betés MT, Súbtil JC, Muñoz-Navas M. Commercially available endoscopy facemasks to prevent aerosolizing spread of droplets during COVID-19 outbreak. Endosc Int Open. 2020 Jun 2;08:E815–6.

4. Marchese M, Capannolo A, Lombardi L, di Carlo M, Marinangeli F, Fusco P. Use of a modified ventilation mask to avoid aerosolizing spread of droplets for short endoscopic procedures during coronavirus COVID-19 outbreak. Gastrointest Endosc. 2020;92:439–40.

5. NHS England. Health Technical Memorandum 03-01 Specialised ventilation for healthcare premises Part A: The concept, design, specification, installation and acceptance testing of healthcare ventilation systems [Internet]. 2021 [cited 2022 May 29]. Available from: https://www.england.nhs.uk/wp-content/uploads/2021/05/HTM0301-PartA-accessible-F6.pdf

6. Chow TT, Yang XY. Ventilation performance in the operating theatre against airborne infection: numerical study on an ultra-clean system. Journal of Hospital Infection. 2005;59:138–47.

7. Rezapoor M, Alvand A, Jacek E, Paziuk T, Maltenfort MG, Parvizi J. Operating room traffic increases aerosolized particles and compromises the air quality: a simulated study. J Arthroplasty. 2018;33:851–5.

8. Gregson FKA, Shrimpton AJ, Hamilton F, Cook TM, Reid JP, Pickering AE, et al. Identification of the source events for aerosol generation during oesophago-gastro-duodenoscopy. Gut. 2022;71:871–8.

9. Gregson FKA, Sheikh S, Archer J, Symons HE, Walker JS, Haddrell AE, et al. Analytical challenges when sampling and characterising exhaled aerosol. Aerosol Science and Technology. 2022;56:160–75.

10. Liu DT, Phillips KM, Speth MM, Besser G, Mueller CA, Sedaghat AR. Portable HEPA purifiers to eliminate airborne SARS-CoV-2: a systematic review. Otolaryngology–Head and Neck Surgery. 2022;166:615–22.

11. Thuresson S, Fraenkel CJ, Sasinovich S, Soldemyr J, Widell A, Medstrand P, et al. Airborne severe acute respiratory syndrome coronavirus 2 (SARS-CoV-2) in hospitals: effects of aerosol-generating procedures, HEPA-filtration units, patient viral load, and physical distance. Clinical Infectious Disease. 2022;75:E89–96.

12. Conway Morris A, Sharrocks K, Bousfield R, Kermack L, Maes M, Higginson E, et al. The removal of airborne severe acute respiratory syndrome coronavirus 2 (SARS-CoV-2) and other microbial bioaerosols by air filtration on coronavirus disease 2019 (COVID-19) surge units. Clinical Infectious Diseases. 2022;75:E97–101.

13. Fujimura H, Nishikawa J, Okamoto T, Goto A, Hamabe K, Sakaida I. Use of portable partitions with high-efficiency particulate air filters in the endoscopy unit. Endosc Int Open. 2021;09:E278–9.

14. Sagami R, Nishikiori H, Sato T, Tsuji H, Ono M, Togo K, et al. Aerosols produced by upper gastrointestinal endoscopy: a quantitative evaluation. American Journal of Gastroenterology. 2021;116:202–5.

15. Morawska L, Johnson GR, Ristovski ZD, Hargreaves M, Mengersen K, Corbett S, et al. Size distribution and sites of origin of droplets expelled from the human respiratory tract during expiratory activities. J Aerosol Sci. 2009;40:256–69.

16. Rayner RK. Bootstrapping p values and power in the first-order autoregression: a Monte-Carlo investigation. Journal of Business & Economic Statistics. 1990;8:251–63.

17. Han ZY, Weng WG, Huang QY. Characterizations of particle size distribution of the droplets exhaled by sneeze. J R Soc Interface. 2013;10:20130560.

18. Tang JW, Nicolle AD, Klettner CA, Pantelic J, Wang L, Suhaimi A bin, et al. Airflow dynamics of human jets: sneezing and breathing - potential sources of infectious aerosols. PLoS One. 2013;8:e59970.

